# Artificial intelligence for personalized management of vestibular schwannoma: A clinical implementation study within a multidisciplinary decision making environment

**DOI:** 10.1101/2023.11.17.23298685

**Authors:** Navodini Wijethilake, Steve Connor, Anna Oviedova, Rebecca Burger, Jeromel De Leon De Sagun, Amanda Hitchings, Ahmed Abougamil, Theofanis Giannis, Christoforos Syrris, Kazumi Chia, Omar Al-Salihi, Rupert Obholzer, Dan Jiang, Eleni Maratos, Sinan Barazi, Nick Thomas, Tom Vercauteren, Jonathan Shapey

## Abstract

**Background:** The management of patients with Vestibular Schwannoma (VS) relies heavily on precise measurements of tumour size and determining growth trends.

**Methods:** In this study, we introduce a novel computer-assisted approach designed to aid clinical decision-making during Multidisciplinary Meetings (MDM) for patients with VS through the provision of automatically generated tumour volume and standard linear measurements. We conducted two simulated MDMs with the same 50 patients evaluated in both cases to compare our proposed approach against the standard process, focusing on its impact on preparation time and decision-making.

**Findings:** Automated reports provided acceptable information in 72% of cases, as assessed by an expert neuroradiologist, while the remaining 28% required some revision with manual feature extraction. The segmentation models used in this report generation task achieved Dice scores of 0.9392 (± 0.0351) for contrast-enhanced T1 and 0.9331 (± 0.0354) for T2 MRI in delineating whole tumor regions. The automated computer-assisted reports that included additional tumour information initially extended the neuro-radiologist’s preparation time for the MDM (2m 54s (± 1m and 22s) per case) compared to the standard preparation time (2m 36s (± 1m and 5s) per case). However, the computer-assisted simulated MDM (CAS-MDM) approach significantly improved MDM efficiency, with shorter discussion times per patient (1m 15s (± 0m and 28s) per case) compared to standard simulated MDM (SS-MDM) (1m 21s (± 0m and 44s) per case).

**Interpretation:** This pilot clinical implementation study highlights the potential benefits of integrating automated measurements into clinical decision-making for VS management. An initial learning curve in interpreting new data measurements is quickly mastered and the enhanced communication of growth patterns and more comprehensive assessments ultimately provides clinicians with the tools to offer patients more personalized care.

**Funding:** N. Wijethilake was supported by the UK Medical Research Council [MR/N013700/1] and the King’s College London MRC Doctoral Training Partnership in Biomedical Sciences. This work was supported by core funding from the Wellcome Trust (203148/Z/16/Z) and EPSRC (NS/A000049/1) and an MRC project grant (MC/PC/180520). TV is also supported by a Medtronic/Royal Academy of Engineering Research Chair (RCSRF1819/7/34).

**Graphical Abstract:** 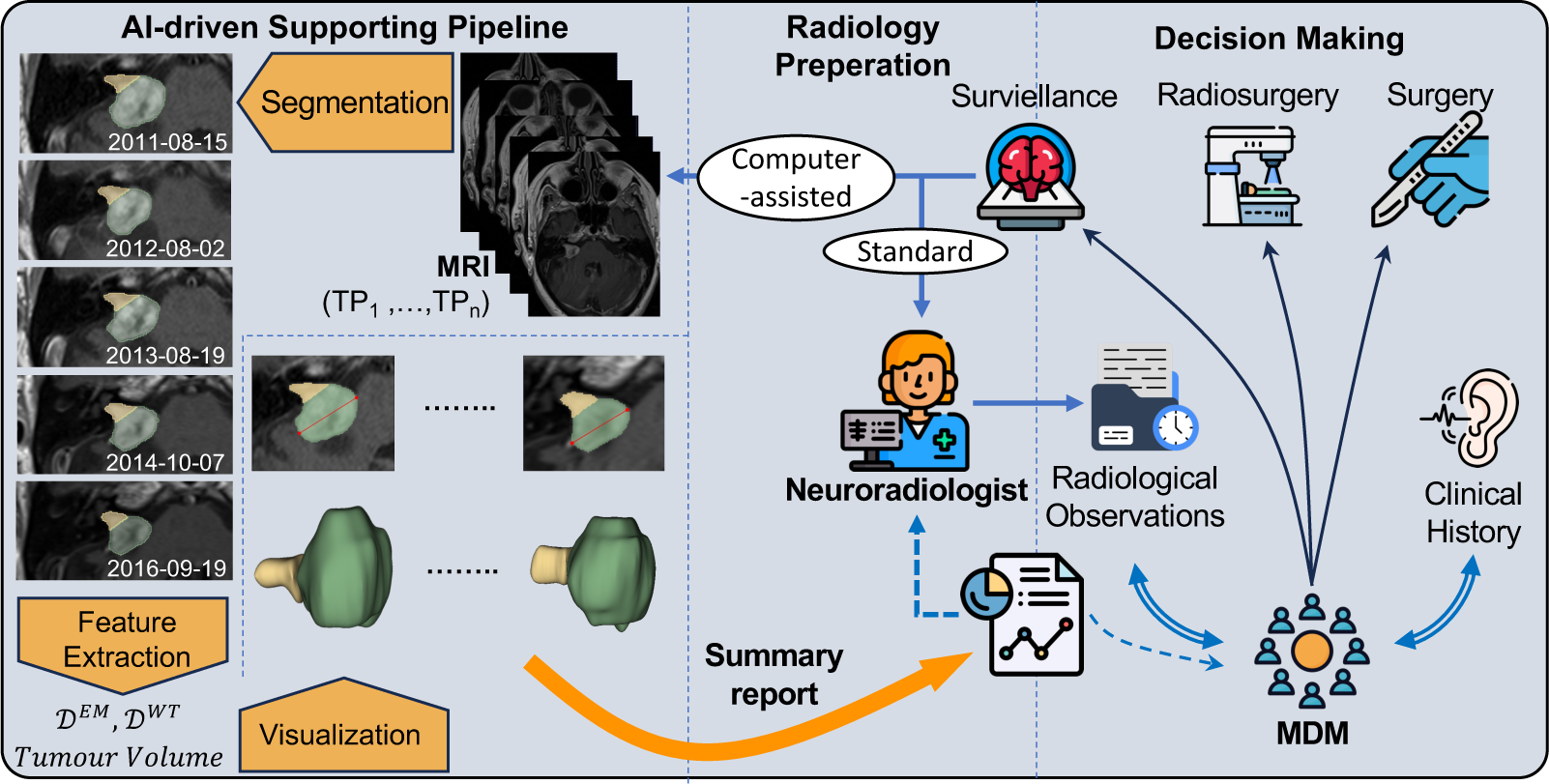

**Highlights:**

- The first study to evaluate the impact of AI assisted reporting in a clinical setting.
- AI generated segmentations can be used to provide a clinical guideline driven report facilitating personalized patient management
- Volumetric tumour measurements provide a more comprehensive assessment of tumour growth.

## 1. Introduction

Vestibular schwannoma (VS) is a benign brain tumour originating from myelinating Schwann cells within the vestibular division of the vestibulocochlear nerve. The incidence rate of VS is rising [1]; in the UK the incidence rate is approximately 2.2 per 100,000 people per year [2] with estimates indicating that approximately 1 in 1000 people will be diagnosed with a VS in their lifetime [3]. The increasing availability and improved quality of Magnetic Resonance Imaging (MRI) has resulted in higher proportion of small asymptomatic tumours now being diagnosed. For smaller tumours, expectant management with lifelong imaging is often advised [4] with patients proceeding to stereotactic radiosurgery (SRS) or conventional open surgery should the tumour demonstrate growth on serial imaging. Even after treatment, patients typically require an extended period of surveillance.

### 1.1. Vestibular schwannoma growth rate criteria

The assessment of VS growth requires a standardized measurement approach [5]. The 2001 Consensus Meeting on Reporting Results in Acoustic Neuroma recommends distinguishing intrameatal and extrameatal portions and measuring the largest extrameatal diameter [6]. When a tumour is entirely intrameatal, this measurement becomes the maximum whole tumour diameter. For VS treatment decisions, both linear and volumetric measurements are applicable. As per the European Academy of Otology & Neuro-Otology (EAONO) position statement, the criteria for significant VS growth include a *>*2 mm increase in diameter, a *>*1.2 cm^3^ volume change, or a *>*20% volume change [7].

### 1.2. Patient pathway

The UK introduced cancer Multi-Disciplinary Meetings (MDMs) to ensure uniform high-quality care for all cancer patients, regardless of origin [8]. Through MDMs the skull base mutlidisciplinary team (MDT) manage complex skull base tumours, such as schwannomas and meningiomas. Core team members include skull base neurosurgeons and otolaryngologists, neuroradiologists, clinical oncologists, histopathologists, clinical nurse specialists, and MDM coordinators. MDMs provide tailored treatment, considering surgery, radiation, and imaging surveillance timing.

### 1.3. Current issues

The manual extraction of linear measurements is a time-consuming process that is susceptible to variation [9]. This introduces interobserver errors that can be significant-even exceeding 2 mm, despite such value being used as a criteria for establishing VS growth. Consequently, these errors may lead to more frequent scans or delayed/unnecessary treatments. Volumetric measurements offer higher sensitivity and precision, but the existing methods, including manual or semi-automated tumour segmentation, are also labour-intensive, lack standardization and are prone to variability and subjectivity [10, 11]. Additionally, the lack of readily available standardized software has hindered their adoption in clinical practice.

### 1.4. Use of Artificial Intelligence in VS

Artificial Intelligence (AI) has emerged as a valuable tool in these studies, contributing to various aspects of VS research. Deep learning and machine learning techniques have been utilized for tumour segmentation [12, 13, 14], growth prediction [15, 16], surgical outcome prediction [17, 18], and Koos grade prediction [19].

Our previous research has shown that AI tools possess the technical capability to fully automate the detection and segmentation of VS [12, 20], and can also delineate the tumour’s intra- and extra-meatal components [14]. These tools formed the basis for automating the extraction process of both linear and volumetric measurements in this study. Nevertheless, while AI has shown promise in VS management, the assessment of its clinical applicability and its impact on decision-making remains an ongoing area of investigation.

### 1.5. Contribution

In this study, we developed an automated imaging biomarker report generator that employs deep learning segmentation and computer algorithms for feature extraction and visualization. Each report included longitudinal measurements of the tumour, axial views of the tumour, and graphical representations and plots illustrating changes in both linear and volume measurements. The main contribution of this study is the assessment of the AI-assisted report in a simulated controlled real-world context. To the best of our knowledge, this is the first study to integrate AI-based outcomes into clinical decision-making for tumour management. The overall process of implementing AI-driven methods in the clinic is visualized in Figure 1 and in this study we evaluate its impact on enhancing the standard clinical management.

**Figure 1:**
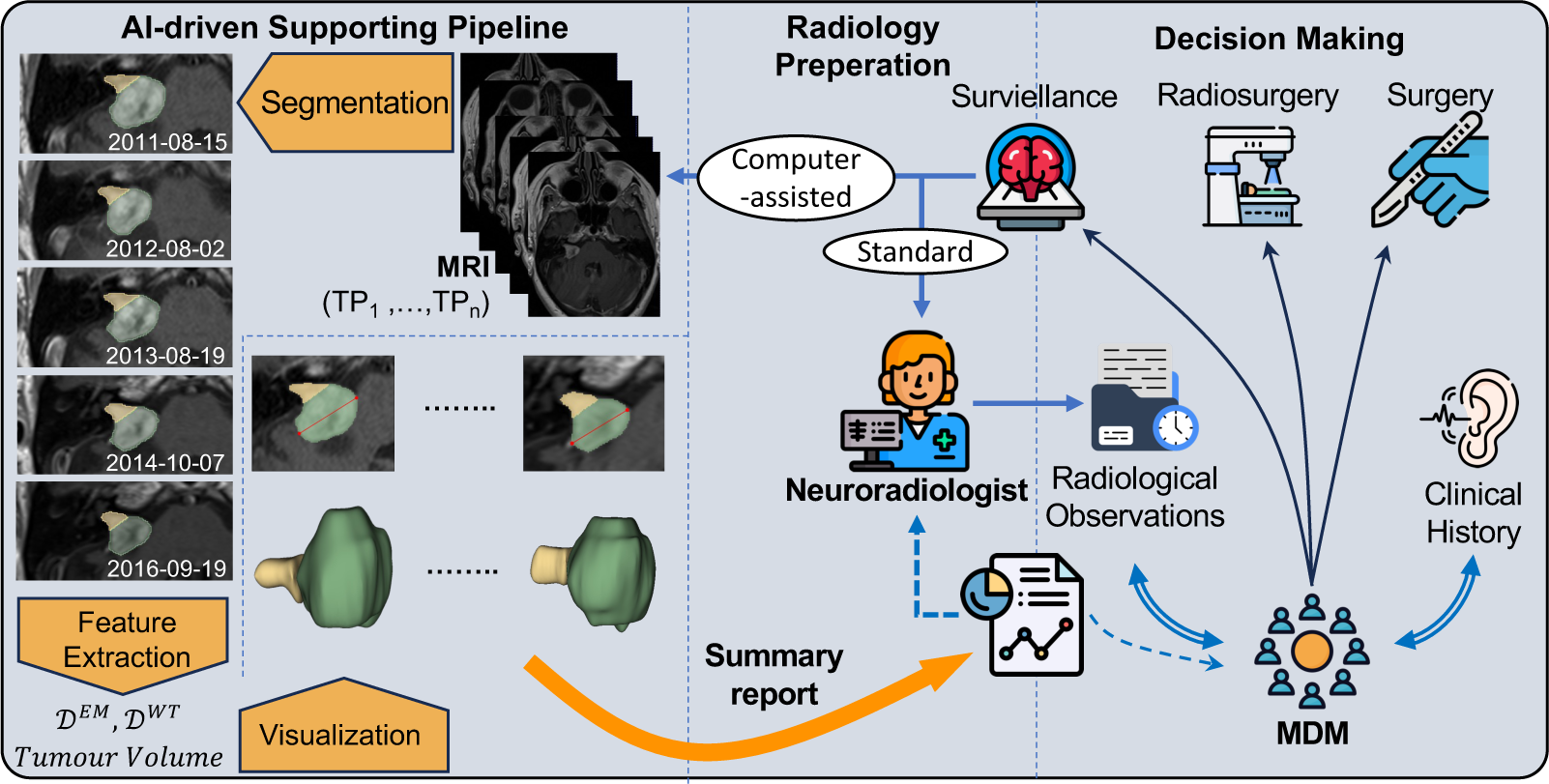
Overview diagram of our clinical implementation study focusing on personalized management of VS.

## 2. Methods

To evaluate the impact on standard clinical practice, two separate simulated MDM sessions involving 50 referred patients were conducted in a controlled setting. The first session adhered to the standard preparation procedure with manual extraction, while the second session introduced the automated imaging biomarker report.

### 2.1. KCH MC-RC dataset

The source cohort for this study consisted of patients referred to King’s College Hospital, London, UK, between December 2009 and September 2012, aged over 18, with unilateral VS, and excluding neurofibromatosis type 2 (NF2).

Fifty patients were randomly selected for our study, with consecutive time points spanning 6 months to 11 years between the first and most recent scan (January 2012 - March 2021). Among these, 8 had prior surgery, 7 received radiosurgery, 1 had both surgery and radiosurgery options, and 34 were under surveillance. The MR imaging included post-contrast T1 (T1C) or T2-weighted imaging. We analyzed 187 sessions across 50 patients, with 145 T1C and 42 T2 scans, including 23 post-operative scans. Table 1 summarizes the demographics of the selected cohort of 50 patients.

**Table 1:** Demographics of the KCH MC-RC patient cohort consists of 50 patients used in the simulated MDM.

| Category | N | Mean | SD |
| --- | --- | --- | --- |
| Age | 50 | 64.62 | 12.84 |
| **Gender** |  |  |  |
| Female | 26 (48%) |  |  |
| Male | 24 (52%) |  |  |
| Monitoring period | 50 | 4.8 (yrs) | 2.74 (yrs) |
| **Standard Tumour size measurement in the index scan** |  |  |  |
| Extrameatal Diameter | 25 (50%) | 18.52 (*mm*) | 9.04 (*mm*) |
| Whole tumour Diameter* | 25 (50%) | 10.36 (*mm*) | 5.84 (*mm*) |
| **Interventional treatment History (prior to the most recent scan)** |  |  |  |
| Surgery | 9 (18%) |  |  |
| SRS | 8 (16%) |  |  |

#### Ethics statement

This study was approved by the NHS Health Research Authority and Research Ethics Committee (18/LO/0532). Because patients were selected retrospectively and the MR images were completely anonymised before analysis, no informed consent was required for the study.

### 2.2. Formation of automated imaging biomarker reports

*Deep learning model development.* Building on the methodology in [14], we adopted a two-stage approach employing the default 3D full-resolution UNet from the nnU-Net framework (referred to as 3D nnU-Net). Additional information regarding the datasets and training procedures can be found in the Supplementary Appendix 1.1 & 1.2. No patient overlap exists between this dataset and the one used for the simulated MDM.

#### Feature extraction and visualization

For the KCH MC-RC dataset, we acquired segmentation masks using the best-performing deep learning models and utilized them in the feature extraction process. The primary linear measurement we extracted for each VS was the maximum tumour diameter. This measurement could be derived from two regions: the entire tumour region (*D^WT^*) and/or the extrameatal region of the tumour (*D^EM^*). When the extrameatal tumour portion was small or negligible, radiologists typically measured the linear dimension for the entire tumour region (combining both intra- and extrameatal regions). More details about the linear features and selection of the appropriate measurement to present are provided in the Supplementary Appendix 1.3. Further, In our previous study [21], we conducted a preliminary evaluation comparing automated and an expert’s manual linear measurements.

Additionally, volume measurements were extracted for the intra- and extra-meatal regions. Visualization of the axial slice with the maximum *D* was done using the 3D Slicer software extension with python [22]. Additionally, 3D volume mask of the tumour was visualized.

#### Automated generation of the imaging biomarker report

Automated report generation followed two key steps: 1) segmentation and 2) feature extraction. For each patient a summary report and an extended version were generated. In the summary report, an MRI axial slice representative of each time point was included, along with the intra-/extra-meatal mask, corresponding extrameatal and whole tumour volumes, and maximum *D* measurement. Volume measurements were not presented if the tumour was present in more than one axial slice. The report included a visual representation of volume and maximum *D* change, using color coding to indicate growth (red), equivocal growth (orange), and no growth (green). Intra-/extra-meatal volumes were displayed in a bar plot, and diameters in a line plot with the x-axis representing time in months. Icons on the graph represented the decisions made after each session, with a guide provided at the bottom of the report. An example report is provided in the Supplementary Appendix 2.2. Additionally, an extended report is generated with all the measurements and 3D visualizations.

### 2.3. Simulated MDMs

#### Preparation for the standard MDM

The neuroradiologist prepared for the SS-MDM under time-constrained conditions to replicate clinical MDM preparation following UK practice guidelines. Linear measurements were extracted from the index, second most recent, and most recent scans for each patient using the Picture Archiving & Communications System (PACS) workstation (Sectra workstation, Sectra AB, Sweden) at King’s College Hospital. For each patient, the neuroradiologist recorded the absolute linear measurements, provided a qualitative description of the tumour, and made judgments regarding tumour growth (change/no change/equivocal) on a structured form. Longitudinal changes were assessed between the index and recent time points and between the second most recent and most recent time points. The starting and finishing times for manual feature extraction for each patient case were recorded. The neuroradiologist’s workload was also assessed using the The NASA Task Load Index (NASA-TLX) scoring system after completing the preparations [23].

#### Execution of the standard MDM

The SS-MDM was conducted with the minimum required attendees, and it took place as a hybrid online-in-person meeting. The simulated skull base MDM included two skull-base neurosurgeons (N.T., J.S.), two clinical oncologists (K.C., O.A.), clinical nurse specialists (A.H., J.D.S.), and three neurosurgical fellows (T.G., A.A., C.S.). Each case was presented to the MDM by the MDM coordinator (A.O). The neuroradiologist (S.C.) then presented the tumour measurements and observations, with the tumour displayed on the PACS system through screen sharing. The starting and finishing times of the MDM were recorded.

#### Preparation for the computer-assisted simulated MDM

The neuroradiologist prepared for the CAS-MDM, 35 days after the SS-MDM preperations, in a time-constrained environment, utilizing the automated report. In this preparation phase, the neuroradiologist evaluated and decided on the acceptance or rejection of automated biomarkers for each patient’s MRI session (time point). This evaluation was based on the segmentation provided by the deep learning model and the automatically extracted features. If a session was rejected, the neuroradiologist manually extracted the linear measurements. The assessments conducted during this process followed the same procedure as in the SS-MDM, utilizing the index, second most-recent, and most recent scans. To reduce the potential for bias, the order of the 50 patient cases was randomized in between the standard and the computer-assisted approach.

#### Execution of the computer-assisted MDM

The CAS-MDM followed the same format and impact assessment criteria as the SS-MDM and was held 35 days after the SS-MDM. Automated reports were distributed to all participants prior to the MDM and the neuroradiologist (S.C.) presented his observations, referencing either linear or volume measurements as necessary. During the CAS-MDM, members also referred to the automated report to assist their decision making.

## 3. Results

### 3.1. Deep learning based segmentation

The performance of the segmentation models during the pre-MDM testing phase are presented in the Supplementary Appendix 2.1.

### 3.2. Neuroradiological preparation for the computer-assisted simulated MDM

In 72% of cases (36 patients), the automatically generated segmentations and linear/volume measurements were accepted and used for assessing tumor growth by the neuroradiologist (Supplementary Appendix 2.2). Out of these 36 patients, 33.33% (12 cases) the neuroradiologist presented the MDM with growth-related observations based on the automated volume measurements provided in the summary report. The remaining cases were assessed using both linear and volume measurements, and in these instances, both linear and volume measurements exhibited consistent changes.

For 16% of patients (8 cases), at most one session among the three sessions had unacceptable segmentation, leading to the rejection of both linear and volume measurements by the neuroradiologist (Supplementary Appendix 2.2). In these cases, the neuroradiologistq manually extracted the linear measurements for the rejected session. The observations, which combined manual and automated measurements, were used to draw conclusions related to tumor growth and presented to the MDM.

In 12% of patients (6 patients), the automated outcomes required complete revision by the neuroradiologist during the preparation of the CAS-MDM. This occurred when at least two sessions, out of the three sessions (index, second most recent, and most recent), had unacceptable segmentations or mismatching linear measurements (Supplementary Appendix 2.2). In such instances, only the neuroradiologist’s manual extractions were used in the MDM.

### 3.3. Simulated MDMs

In the SS-MDM, 7/50 patients were referred for active treatment discussions with patients, including 5 for SRS and 2 for surgery. 4 patients were discharged, and the rest were referred to be placed under surveillance. The patients under surveillance will be rescanned in 6 months (2 patients), 1 year (9 patients), 2 years (16 patients), 2-3 years (1 patient), 3 years (7 patients), 4 years (1 patient), and 5 years (3 patients).

In the CAS-MDM, 6 patients were referred for active treatment discussions, including 5 for SRS and 1 for surgery. 6 patients were discharged, while the rest were recommended to remain under surveillance. The patients under surveillance will be rescanned in 6 months (3 patients), 1 year (9 patients), 2 years (11 patients), 2-3 years (4 patients), 3 years (9 patients), 3-4 years (1 patient), and 5 years (1 patient).

The average times for preparation and MDM for both standard and computer-assisted approaches are presented in Table 2 with the distribution visualized in Figure 2 (B).

**Figure 2:**
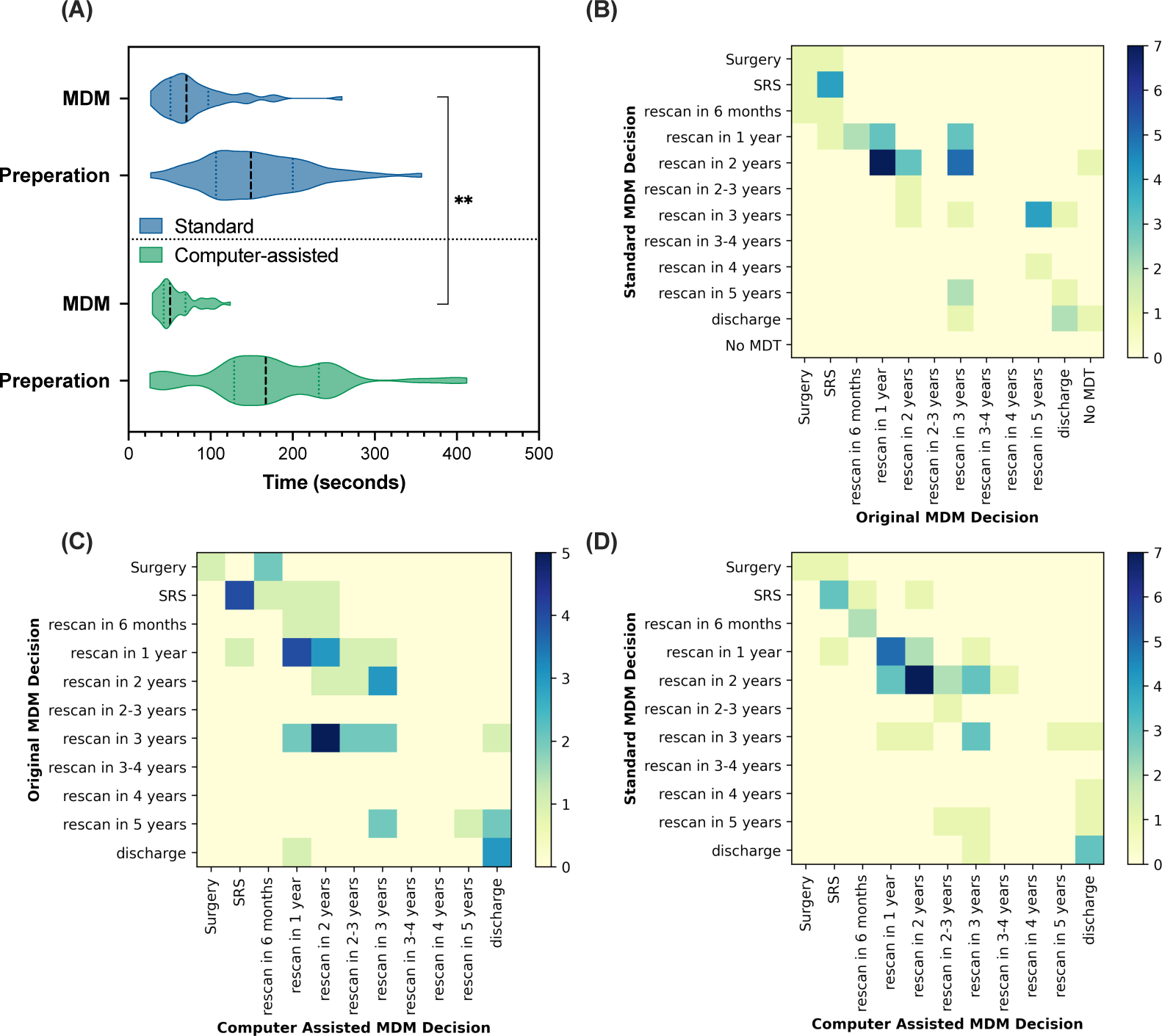
(A) Comparison of the timings of radiology preparation and MDM for standard and computer-assisted approaches. (B) Confusion matrix between the SS-MDM decisions and the (actual) original MDM decision (C) Confusion matrix between the (actual) original MDM decisions and the CAS-MDM decisions. (D) Confusion matrix between the SS-MDM and the CAS-MDM decisions.

**Table 2:** Comparison of Preparation and MDM Process Durations in SS-MDM and CAS-MDM Sessions.

|  |  | Average time per case | Total time |
| --- | --- | --- | --- |
| Standard | Preparation | 2m 36s (± 1m and 5s) | 2h 7m 30s |
| | MDM | 1m 21s (± 0m and 44s) | 1h 2m 36s |
| Computer-assisted | Preparation | 2m 54s (± 1m and 22s) | 2h 25m 24s |
| | MDM | 1m 15s (± 0m and 28s) | 48m 24s |

## 4. Discussion

In this study, we introduce a novel approach, CAS-MDM, that integrates automated imaging biomarkers reports into VS clinical management. We validate this concept in a controlled simulated clinical setting, involving experts from multiple disciplines. Through deep learning based automated segmentation, we provide highly sensitive volume measurements, crucial for analyzing VS growth. This approach has the potential to streamline decision-making, enhancing clinical efficiency, and offers potential for more personalized patient care. The use of automated reporting resulted in a substantial improvement in clinical efficiency and the potential to enhance patient management quality.

To the best of our knowledge, our research represents the first clinical deployment of an AI supported report for tumour management, where we examine its affect on clinical workflow and decision-making process. Hawkins et al. [24] recently implemented a machine learning-based clinical workflow to perform segmentation, volumetric calculations, and generate reports using longitudinal MRI scans for low-grade glioma. However, their study did not explore how this technology could be integrated into clinical decision-making.

### 4.1. Neuroradiological preparation

During preparations for the CAS-MDM, the neuroradiologist manually extracted linear measurements when the whole tumor segmentation was unacceptable at any of the three time points (index, second most recent, and most recent).

We identified 4 scenarios contributing to faulty segmentations in Figure 3. Figure 3 (A) visualizes a session with an over-segmentation of a small tumor, while Figure 3 (B) and Figure 3 (C) depict instances of under-segmentation and non-recognition by the segmentation model, respectively. As a result, the neuroradiologist manually extracted the linear measurements for these cases. Additionally, Figure 3 (D) is an example of a session that was over-segmented, and it did not provide an acceptable intra-/extra-meatal boundary.

**Figure 3:**
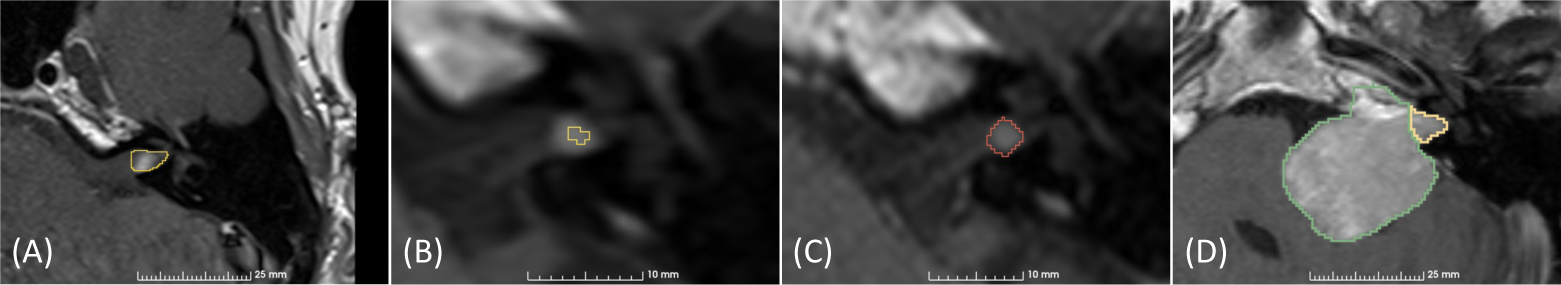
Examples of deep learning generated segmentations rejected by the neuroradiologist. (A) Over segmented slice. (B) Under segmented slice. (C) Missed tumour detection. The manual annotation of the tumour region is shown in red. (D) Over segmented and the incorrect intra-/extra-meatal boundary.

During the SS-MDM preparations, the neuroradiologist chose between *D^WT^* and *D^EM^*, based on whether the maximum extrameatal dimension appeared larger than the porus on axial images. If the neuroradiologist was seeking serial comparable dimensions and the initial tumour is intrameatal, then the later measurement should also consider the entire tumour, even if it has developed a significant extrameatal component. This posed a significant challenge in the context of the automated imaging biomarker report, as we selected *D^EM^* according to our pseudo code Supplementary Appendix 1.3. Consequently, the neuroradiologist identified this as an inconsistency in the automated imaging biomarker report and relied on volume measurements.

Furthermore, for several cases, the whole tumour volume tended to provide more reliable observations on growth, as shown in Figure 4. Figure 4 (A) displays post-operative yearly scans between 2011 and 2013, along with corresponding *D^WT^* and whole tumour volume measurements. The change between the second and third time points indicates definitive growth (a change in volume of over 20%), while the change in linear measurement (*D^WT^*) suggests equivocal growth.

**Figure 4:**
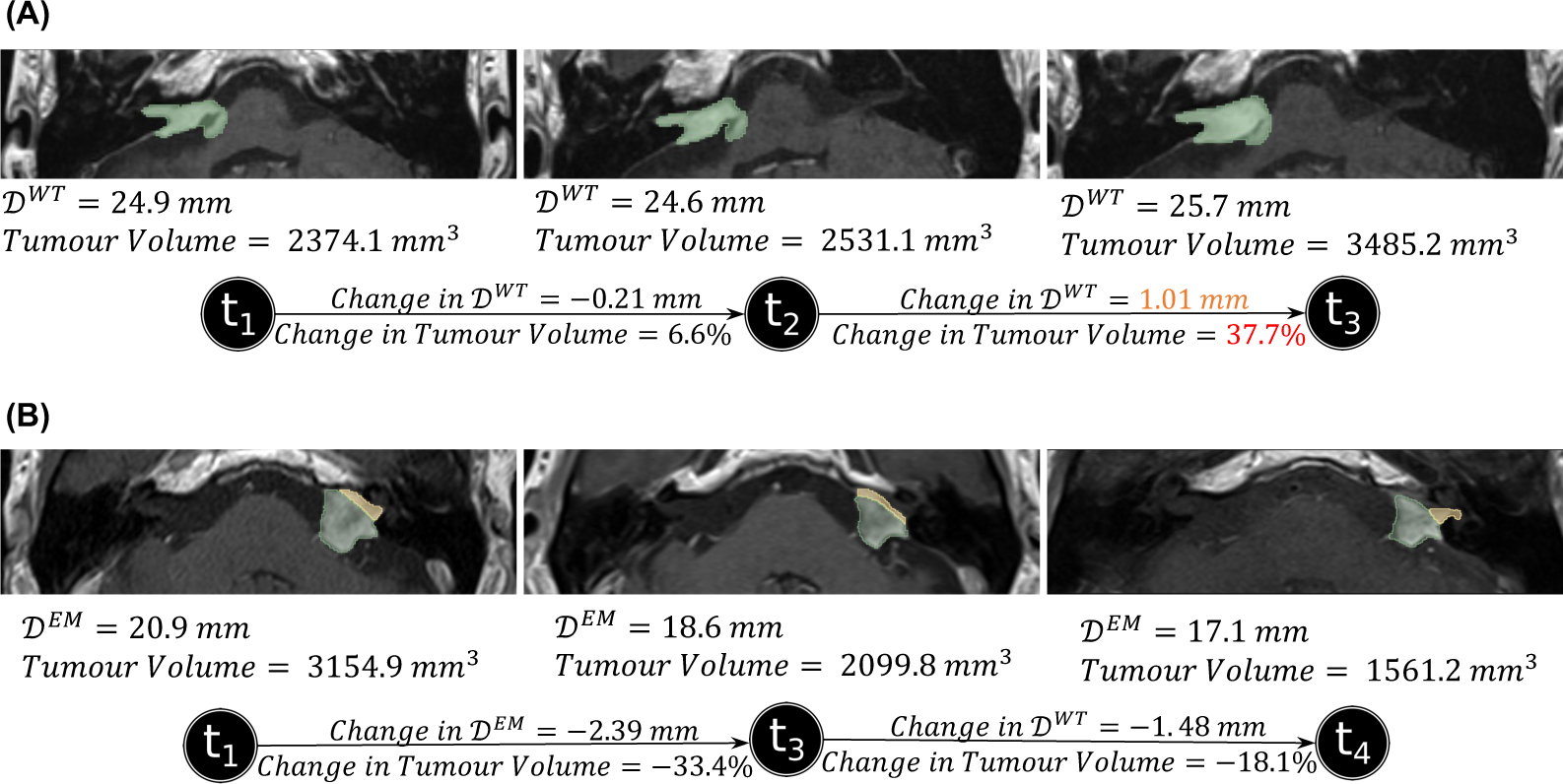
Two patient cases that depicts the importance of volume measurements. (A) A patient case where the volume measurement change presents a definitive growth in contrast to the linear measurement. (B) A patient case with an inconsistent and irregular intra-/extra-meatal boundary, for which the tumour volume is more reliable.

Inconsistency in the boundary between the longitudinal scans was another reason for choosing whole tumour volume for some patients. Figure 4 (B) presents longitudinal scans of a post-SRS patient from 2012, 2016, and 2019 who had undergone SRS in 2005. Due to the irregular shape of the tumour, the separation of the intra-/extra-meatal regions does not accurately depict the tumour’s behavior over time. These observations were highlighted by the neuroradiologist during the preparations and also when presenting to the MDM.

Our results demonstrated that the average preparation time for the CAS-MDM was longer than for the SS-MDM which was surprising. However, this was partly due to the learning curve experienced by the neuroradiologist, which might have been mitigated through training on a separate trial cohort. Furthermore, in the CAS-MDM, the neuroradiologist had to assess both the validity of segmentations and linear/volume measurements, which was different to the SS-MDM preparations that exclusively utilized linear measurements. Both the SS-MDM and CAS-MDM preparations placed a medium workload on the neuroradiologist according the NASA-TLX scoring interpretation however information overload on the summary report may have also contributed to the extended preparation time.

The neuroradiologist’s preparation may be negatively affected by poor performance of the segmentation model. This may be a result of various factors including limited availability of the desired MRI sequences, non-standard MRI acquisition, case complexity, tumour heterogeneity, the presence of very small tumours or residual tumour following surgery. Better automated segmentation results are likely to be achieved through the standardisation of MRI sequences used to image VS. Nevertheless, minor adjustments will almost certainly be required in some difficult cases so an integrated interactive segmentation module, allowing the neuroradiologist to make real-time adjustments in the segmentation masks, should be developed before such technology can be fully integrated into the routine clinical workflow. Integrating such a module into PACS would further reduce the cognitive load of processing/visualizing various metrics, enabling the neuroradiologist to filter and focus on a specific preferred metric.

### 4.2. Observations from the simulated MDMs

The total number of discharges were higher in the CAS-MDM (6 patients) compared to the SS-MDM (4 patients). The MDT agreed to discharge three of the same patients in both MDMs. One patient discharged from the SS-MDM was not discharged from the CAS-MDM; three additional patients were discharged from the CAS-MDM. Discharged patients typically included those in old age (¿80 years) with small stable tumours. Figure 2 (C) summarizes the similarities and differences between the SS-MDM and the CAS-MDM.

In the SS-MDM, of 39 patients for whom surveillance imaging was recommended, the rescan interval increased for 11 patients (28.2%) decreased for 8 patients(20.5%), and remained unchanged for 16 patients (41%) as compared with the MDTs original clinical decision. Figure 2 (B) illustrates how decisions taken in a SS-MDM can change over time based on the original decisions made and the simulated SS-MDM decisions for the 50 patients. Patient management may have changed since the patient’s original clinical MDM due to evolving clinical management and variations in the composition of MDM participants together with their individual biases toward different treatments.

The use of computer-assisted reporting enabled the MDT to offer patients a personalized management approach beyond standard surveillance protocols. Decision-making in both SS-MDM and CAS-MDM was primarily based on tumour size measurements. However, other factors such as the patient’s age and treatment history significantly influenced the decisions.

The team’s oncologists particularly valued having whole tumour volume measurements available as this is the metrics used when delivering radiotherapy-not just the extrameatal portion. Additionally, graphical representations illustrating growth trends over the patients whole surveillance programme were beneficial in determining whether to extend the interval between surveillance scans.

From the neuroradiologist’s perspective, the automated imaging biomarker report, with its multiple inputs (linear and volume measurements for each imaging timepoint), was more challenging to interpret compared to the standard approach, where only index, second most recent, and most recent imaging were used. Consequently, in the CAS-MDM, observations were limited to the same three sessions to reduce complexity. However, the communication of trends across multiple timepoints proved immensely beneficial, and the inclusion of additional volume data played a vital role in enhancing the precision of interval change assessments. Most importantly, the neuroradiologist highlighted the importance of keeping an expert human in the loop for the absence of thorough data verification may lead to clinicians misinterpreting critical information.

### 4.3. Automated patient-centric report

We organized a Patient Involvement Group (PPI) focus group in partnership with the British Acoustic Neuroma Association (BANA), to explore how this technology could be used to provide patients with graphical information concerning their treatment. We selected 12 individuals from a pool of over 200 interested patients across the UK, ensuring diversity in age, gender, and treatment experiences. This session was conducted online for accessibility.

This PPI session provided valuable insights into patient preferences for automated reports, emphasizing the necessity of visual and personalized formats. Patients highlighted the significance of clarity and simplicity in reporting. An artistic representation (Figure 5) visually depicted the event, referencing the key components highlighted by the patients.

**Figure 5:**
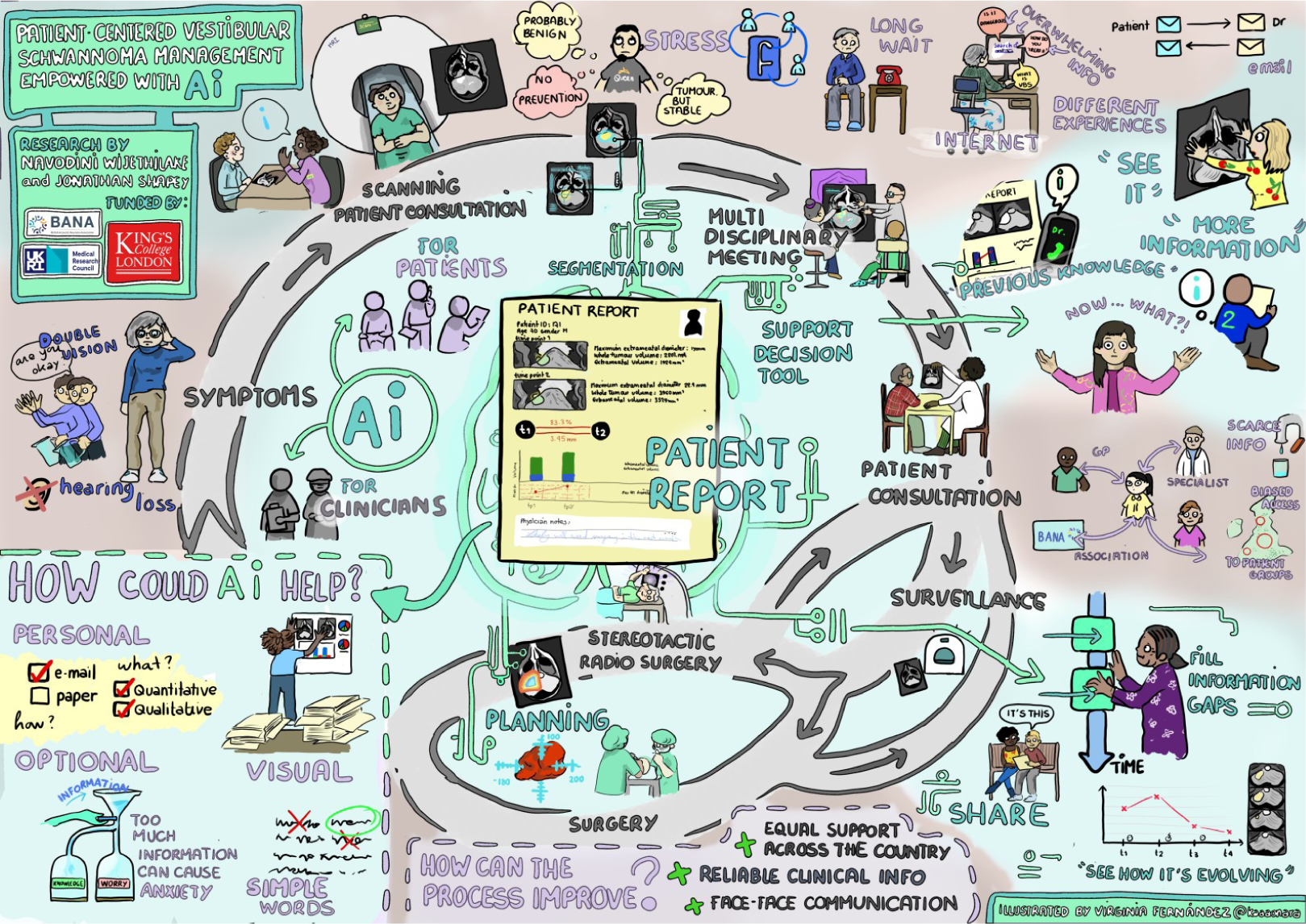
Visual illustration from the patient involvement session.

Our focus group highlighted how this report could be extended for use in combined MDM clinics, as discussed in [25], where patients can access their personalized reports. This could improve efficiency, patient-reported outcomes, decision transparency, and patient mental health.

## 5. Conclusion

This is first study to report the clinical deployment of an AI supported reporting tool for tumour management. We introduced computer-assisted automated reporting exploiting deep learning-based segmentation to aid vestibular schwannoma management and evaluated its impact in a simulated clinical environment. This work demonstrated significant improvements in clinical efficiency and highlighted the potential computer-assisted reporting could have in delivering more personalized patient care.

## Data Availability

Data is partially available on TCIA. The rest will be released on TCIA in the future.

https://wiki.cancerimagingarchive.net/pages/viewpage.action?pageId=157287455#1572874559ba0f6d7166845d88ea4d96f32f0c097

